# Knowledge, Attitudes and Practices of Barangay Health Workers in the Top 5 COVID-19 Affected Barangays in Cebu City Towards Preventing COVID-19 Transmission

**DOI:** 10.1101/2025.04.22.25326079

**Authors:** Elijah Mariano L. Arias, Leah Vanessa Baguio, Sharyz Dane T. Cabalhin, Jennilyn Christy L. Cordero, Ramon Benedicto C. Farrarons, Lemuel Hope S. Galindo, Glydenne Glaire P. Gayam, Ma. Katrina Ada F. Mabelin, Mariadel U. Matute, Tricia Marie A. Niere, Kristina Jean A. Pragados

## Abstract

**Introduction:** COVID-19 has impacted almost the entirety of the world. In the Philippines, the disease has reached a total of 1,269,465 cases and 21,898 deaths as of June 07, 2021.The COVID-19 prevention strategy of the country places Barangay Health Workers (BHWs) at its core. Their basic knowledge, attitudes, and practices (KAP) on COVID-19 control is necessary to allow efficient public health response.

**Objective:** To assess the KAP of BHWs on preventing transmission and exposure to COVID-19.

**Methodology:** The study used a descriptive-correlational research design. A KAP questionnaire was formulated and the proposal was then submitted to the Cebu Institute of Medicine Ethics Review Board. Letters, informed consent forms and questionnaires were then sent to the participating barangays.

**Data Analysis:** A scoring mechanism was applied to the total score of each participant in each category (KAP) and weighted mean was calculated. Lastly, the significant relationship between the KAP of BHWs in the prevention of COVID-19 transmission was calculated and compared.

**Results and Discussion:** Most of the BHWs were classified as knowledgeable (66.10%). None of them scored average to no knowledge at all. In the attitude domain, a factor average of 4.15 was computed which was translated as favorable, and practice classified as Always with an average of 4.43. Using Pearson rho-correlation analysis, results show that knowledge does not necessarily affect attitude (p-value >0.05) but significantly contributes to improving attitude and frequency of practices. Likewise, attitude significantly (p-value < 0.05) induced a positive impact on the practices. Regression Analysis shows that a unit increase in knowledge increases the frequency of practice by 0.7 units and every unit increase of attitude contributes 0.21 to the frequency of practices. Coefficient of variation indicates that 19.2%-21.9% of the total variations of practice can be explained by the variations of knowledge and attitude.

**Conclusion:** The BHWs were found to be knowledgeable in COVID-19 prevention, and their knowledge significantly improved their attitude and frequency of practices. Similarly, their attitude induced a positive impact on their practices.

## INTRODUCTION

Coronavirus disease 2019 (COVID-19) has affected most, if not all, countries in the world. The transmission of severe acute respiratory syndrome coronavirus 2 (SARS-CoV-2) has risen exponentially despite the efforts of several countries to regulate the spread of the virus. As of June 07, 2021, a year and six months since the start of the pandemic, a total of 173,005,553 cases, and 3,727,605 deaths worldwide have been recorded [1]. It is known that the virus is transmitted through direct contact or through droplets that are spread by coughing or sneezing but there have been reports that airborne transmission is also plausible [2][3]. Bearing this in mind, there have been several guidelines from the World Health Organization (WHO) to prevent exposure from the virus with the use of personal protective equipment (PPE) including face masks, frequent hand washing and social distancing [4].

Several countries have succeeded in suppressing the spread of the virus while others have performed poorly in this pandemic. In the United States, there have been reports of individuals that contest the mandatory wearing of masks in public and currently, they hold the highest number of confirmed cases and deaths [5]. There has also been a resurgence of cases seen in European countries and a new variant that has warranted a second lockdown [6][7]. In Asia, Singapore has proven their competence in handling an outbreak while their neighboring countries have struggled due to an unprepared health care system and low public awareness [8].

In the Philippines, despite the attempts to control the spread of coronavirus through repeated community quarantine and the protocols implemented, the number of confirmed cases are still high with a total of 1,269,478 cases and 21,898 deaths. [1][9]. Initially, the COVID-19 cases were clustered in Luzon which have spread across the country and affected numerous cities and provinces including Cebu. Cebu became the epicenter of COVID-19 for weeks and factors including multiple high-density urban-poor neighborhoods, delayed response, lenient contact tracing, insufficient swab testing, and limited information about the virus have been implicated in the surge of cases in the area [10]. Furthermore, there have been multiple reports of mass gatherings and protocol violations which increased the viral transmission [11].

The unforeseen surge of COVID-19 cases in several barangays in Cebu City warranted an immediate response from the local government of Cebu. They responded with stringent implementation of the Inter-Agency Task Force (IATF) guidelines, strict border controls, lockdown and mass testing of the affected barangays [12]. With the tertiary hospitals becoming saturated with the influx of COVID-19 patients, it became overwhelming to cater to all suspected COVID-19 cases and those asymptomatic patients. It became clear that the prevention and early detection of the virus must begin at the community level.

BHWs in coordination with the other sectors play a vital role in controlling the transmission of the virus and reducing the risk of outbreaks in the community [13]. They provide basic primary health care such as medical care, information, counseling and referral [14]. The importance of the adequacy of the KAP of health workers about the ongoing pandemic cannot be overly emphasized. This is of particular interest since it may impede public health response, specifically, ignorance and confusion of basic information such as transmission and exposure, places the people at risk of infection [15][16][17]. It is critical for the planning and implementation of effective pandemic response to understand the public perceptions and responses to the pandemic.

This study aims to assess the KAP of BHWs on preventing transmission and exposure to COVID-19. Specifically, this study aims:

1. To determine the demographic profile of the respondents in terms of age, sex, educational attainment, and monthly salary;
2. To identify the level of knowledge among BHWs in the top 5 affected barangays in Cebu towards preventing COVID-19 transmission;
3. To determine the behavior and attitude of BHWs towards preventing the transmission of COVID-19 in Cebu City;
4. To identify how the knowledge and attitudes are applied through the actions of BHWs in preventing the transmission of COVID-19; and
5. To know the significant relationship between the KAP of BHWs and the prevention of transmission in the 5 selected barangays in Cebu City.

## METHODOLOGY

### Study Design

A descriptive-correlational research design approach was used to achieve the objectives of the study. After quantifying and analyzing the KAP score, multiple characteristics such as demographics affecting the score were explored to establish a relationship. The survey was deemed appropriate for this study since it allowed assessment of the participants with minimal contact to limit the exposure to COVID-19. Furthermore, a purposive sampling was used wherein the respondents already identified the participants of the study, particularly the BHWs.

### Study Setting and Time Period

The data were collected from the top 5 COVID-19 affected barangays of Cebu City based on the DOH region 7 COVID-19 tracker as of January 28, 2021, namely Guadalupe, Mambaling, Lahug, Labangon, and Apas[18]. Placing the focus into these 5 most affected areas allowed us to take a look into specific behaviors which eventually led to the spike in cases that were nearly controlled. Fifteen questionnaires were personally handed and distributed to the barangay hall of each of the said barangays of Cebu City during March 2021 since there was a predicted surge secondary to the holiday’s ease of social distancing protocols.

### Study Population

The study population included the BHWs from the top 5 COVID-19 affected barangays of Cebu City who were involved in assessment, contact tracing, and exposure to different patients through their line of work. BHWs not working in their respective locale at the time of the COVID-19 outbreak were not included in the study.

### Sampling Size Calculation

This study utilized purposive sampling and the participants were limited only to BHWs of the top 5 COVID-19 affected barangays in Cebu city. The total number of BHWs for all 5 barangays is 75. Out of 75, a sample size of 67 was calculated at 95% confidence level and 4% confidence interval.

### Data Collection Process

Once the questionnaire had undergone content validity and had been approved, an informed consent was attached together with the questionnaire. Letters addressed to the city health officer were sent, requesting for permission to allow the researchers to conduct a survey among BHWs. The city health officer and barangay captains of each barangay also checked the contents of the questionnaires. A pilot test was conducted in Barangay Bacayan before investing in a full-scale research survey. After obtaining consent for participation in the study, the questionnaire was handed personally. The questionnaire contained KAP questions related to COVID-19 transmission to collect the needed data.

### Data Collection Tools and Data Analysis

A survey method instrument with integrated KAP questions related to COVID -19 was used in this study.

KAP assessment helped the researchers generate data that may be used to deepen the understanding of commonly known information, attitudes and factors that influence behavior. Questionnaire was obtained originally from a study by Azlan et al., and Erfani et al. with few modifications and additions [19][20]. The modification of the questions were done based on the IATF health protocols in the Philippines.

The survey was composed of 1) demographics, which includes sociodemographics, gender, age, other occupations, level of education, and household income; 2) knowledge about COVID-19; 3) Attitudes towards COVID-19; 4) Practices towards COVID-19. The survey was translated to the Cebuano dialect. A backward translation approach and consultation of a bilingual expert was accomplished in order to assure linguistic and conceptual equivalence.

The survey items under knowledge included 13 questions mostly clinical presentation, transmission routes, and prevention and control. Participants were given “true” or “false” response options to these items. A scoring mechanism was applied to the total score of each participant classified as:

In measuring attitudes towards COVID-19, the participants were asked 11 questions and response options based on a 5-point likert scale ranging from very favorable to very unfavorable attitude. It determined the behavior and attitude of BHWs towards preventing the transmission of COVID-19 in Cebu City. The researchers asked 19 questions about practices and response and options are based on a 5-point likert scale. The items included the IATF’s guidelines and protocols in public places as well as the vaccine protocols yet to be released. In order to rank the level of attitude, the total attitude score was classified as:

For measuring practices towards COVID-19, 19 questions were asked and a 5-point likert scale was used ranging from always to never. Total practices score were classified according to:

Data collection was performed by reaching out to the different barangays (respondents) and sending out paper survey questionnaires.

### Validity

The research instrument was patterned after the study conducted by Azlan AA et al. entitled, Public KAP towards COVID-19: A cross-sectional study in Malaysia. Their instrument was ultimately adapted from a survey that has been previously tested and utilized in China. A COVID-19 knowledge questionnaire was developed by the authors and was based on guidelines for clinical and community management of COVID-19 by the National Health Commission of the People’s Republic of China. In order to adapt to the local setting, the modified questionnaire had a total of 13 knowledge, 11 attitude, and 19 practice questions in comparison to the K-12 A-2 P -2 items of the cited study. This was modified in relation to IATF national guidelines, new medical updates and adapting protocols imposed. There are formidable changes on the approach COVID-19 and it is important to assess them to provide accurate descriptions of the level of KAP of the BHWs.

### Reliability

A pilot study was done prior to the actual data collection with one barangay not included in the study. At least ten samples from BHWs were collected. The data was tallied and analyzed using the Cronbach alpha test.

Consistency of the data was tested using Test-Retest Reliability wherein the questionnaires were given twice to the same population at different times to see if the scores are the same. The two scores are then correlated. A Test-retest reliability coefficient of 0.9% would indicate a very high correlation thereby with good reliability and a value of 10% would indicate a very low correlation, suggesting poor reliability of the instrument and the need for revision or improvement of the questionnaire.

### Plan for Analysis

After data collection, it was organized, tabulated and interpreted. T-test was used for the statistical analysis of the data. Statistical Package for the Social Sciences (SPSS) Version 16 was used to calculate the mean, standard deviation and p-value.

### Statistical Treatment

After data collection, the data were organized, tabulated and interpreted. Frequency and simple percentage was used to determine the demographic profile of the respondents in terms of age, sex, educational attainment and monthly salary.

For the data collected from the knowledge portion, the number of respondents who answered “true” were counted, and likewise, for those who answered “no” to each question per barangay. For the data collected on the attitude and practices portion, wherein a 5-point Likert scale is used, weighted mean was utilized to determine the attitude and practices of the BHWs of each barangay in the prevention of COVID-19 transmission. Pearson r for the correlation of the factors, with p-value < 0.05 as significantly, was used to determine the significant relationship between the KAP of BHWs in the prevention of COVID-19 transmission. A pilot test of at least 10 dry run samples was done, and Cronbach’s alpha was used to evaluate it.

### Ethical Considerations

A protocol was submitted to the Institutional Review Board (IRB) of Cebu Institute of Medicine (CIM) for approval. The disclosure of personal information such as the age, gender, highest level of education and monthly salary of the participant was asked. The risks include the possible exposure to individuals not from the barangay and the questionnaires serve as a vector of the virus if not properly disinfected prior to distribution and upon collection. The participants were informed of the benefits of this study, which is to provide an overview of the COVID-19 response at the barangay level, which may aid in the control of COVID-19 transmission in the community. An informed consent was given prior to participation, and it emphasized the voluntary participation of the respondent confidentiality of data, and stated the potential risks and benefits of the study. Only the tabulated data was presented for the benefit of the study and to answer the objectives. The original forms containing the responses were shredded and disposed properly.

## RESULTS AND DISCUSSION

A survey was conducted to the consenting BHWs in the Top 5 COVID-19 Affected Barangays in Cebu City towards preventing COVID-19 transmission. Out of the 59 total respondents, 11 (19%) came from Apas, 13 (22%) from Guadalupe, 15 (25%) from Labangon, and 10 (17%) from Lahug and Mambaling. The data were then tabulated, analyzed and interpreted.

Table 5 shows the distribution of the respondents in terms of their demographic profile. It can be verified from the table that most of the BHWs (47.46%) who participated were already in their 49 to 58 years of age, all were females, majority were high schools graduates (54.24%), and nearly all claimed they are earning within an income range of 5,000-10,000 every month (96.61%).

**Table 1.** Number of Cases from April 2020-February 2021 and number of assigned health care workers in the top 5 COVID-affected barangays in Cebu City.

| Top 5<br>COVID-affected<br>Barangays | Number of cases (April 2020 - Feb 2021) |  | Number of<br>assigned Health<br>Care Workers |
| --- | --- | --- | --- |
|  | Total confirmed cases | Total confirmed deaths |  |
| Guadalupe | 976 | 44 | 15 |
| Mambaling | 912 | 35 | 15 |
| Lahug | 693 | 26 | 15 |
| Labangon | 638 | 26 | 15 |
| Apas | 571 | 12 | 15 |

**Table 2.** Knowledge score rating scale for BHW Knowledge on COVID-19 transmission.

| **Knowledge score** | **Verbal description** | **Interpretation** |
| --- | --- | --- |
| 13 | Very Knowledgeable | Has excellent knowledge about COVID-19 transmission |
| 9-12 | Knowledgeable | Has adequate knowledge about COVID-19 transmission |
| 5-8 | Average | Has moderate knowledge about COVID-19 transmission |
| 1-4 | Less knowledgeable | Has limited knowledge about COVID-19 transmission |
| 0 | No knowledge at all | Has poor knowledge about COVID-19 transmission |

**Table 3.**
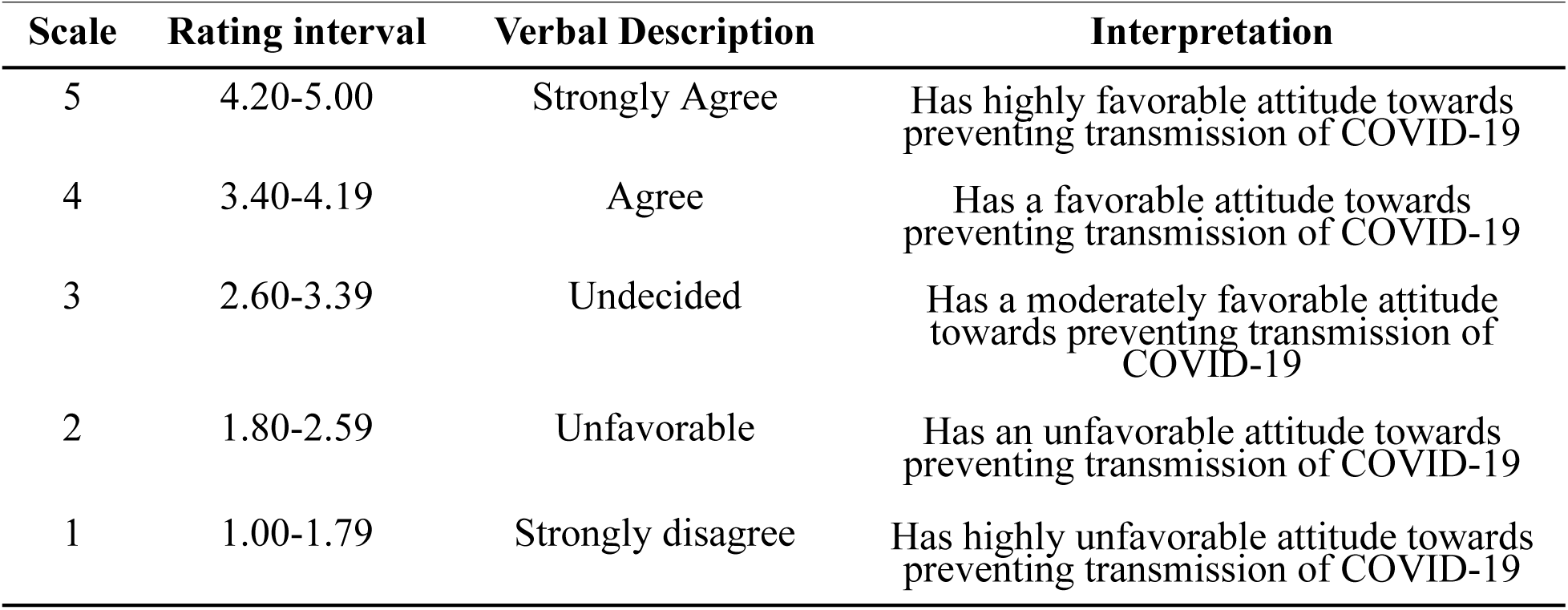
5-Point Rating Scale for Attitude on BHW prevention of COVID-19 transmission.

| **Scale** | **Rating interval** | **Verbal Description** | **Interpretation** |
| --- | --- | --- | --- |
| 5 | 4.20-5.00 | Strongly Agree | Has highly favorable attitude towards preventing transmission of COVID-19 |
| 4 | 3.40-4.19 | Agree | Has a favorable attitude towards preventing transmission of COVID-19 |
| 3 | 2.60-3.39 | Undecided | Has a moderately favorable attitude towards preventing transmission of COVID-19 |
| 2 | 1.80-2.59 | Unfavorable | Has an unfavorable attitude towards preventing transmission of COVID-19 |
| 1 | 1.00-1.79 | Strongly disagree | Has highly unfavorable attitude towards preventing transmission of COVID-19 |

**Table 4.** 5-Point Rating Scale for Practices on BHW prevention of COVID-19 transmission.

| Scale | Rating interval | Verbal Description | Interpretation |
| --- | --- | --- | --- |
| 5 | 4.20-5.00 | Always | Constantly practices the prevention of COVID-19 transmission |
| 4 | 3.40-4.19 | Often | Mostly practices the prevention of COVID-19 transmission |
| 3 | 2.60-3.39 | Sometimes | Sometimes practices the prevention of COVID-19 transmission |
| 2 | 1.80-2.59 | Rarely | Seldomly practice the prevention of COVID-19 transmission |
| 1 | 1.00-1.79 | Never | Does not practice the prevention of COVID-19 transmission |

**Table 5.** Demographic Profile of the BHWs (n=59)

| **Age Range (years)** | **f** | **%** |
| --- | --- | --- |
| 28 to 38 | 10 | 16.95 |
| 39 to 48 | 15 | 25.42 |
| 49 to 58 | 28 | 47.46 |
| 59 to 68 | 6 | 10.17 |
| **Sex** |  |  |
| Female | 59 | 100.00 |
| Male | 0 | 0.00 |
| **Highest Educational Attainment** |  |  |
| College Graduate | 23 | 38.98 |
| College Level | 3 | 5.08 |
| High School Graduate | 32 | 54.24 |
| Vocational | 1 | 1.69 |
| **Monthly Income (in pesos)** |  |  |
| < 5000 | 2 | 3.39 |
| 5,000 to 10,000 | 57 | 96.61 |

Table 6 shows the level of knowledge of the BHWs on different measures to prevent COVID-19 transmission in their respective areas. Overall, 89.31% of the BHWs answered correctly on the different items included in this domain. The findings of this study on the aspect of knowledge on COVID-19 is comparable to similar studies in Nepal and China which showed 81.5% and 89% of the healthcare workers having sufficient knowledge, respectively [21][22]. In fact, it is easy to validate that all of the BHWs knew that “*The main clinical symptoms of COVID-19 are fever, loss of taste, fatigue, dry cough and body aches*” and at the same time “*to encourage symptomatic individuals to stay at home unless there is a pressing need to go to a health facility for medical consultation*” as the best thing to do being a public healthcare worker. In comparison, a study on the KSA of COVID-19 among income-poor households in the Philippines showed that 89.5% of the respondents identified that coughing and sneezing were primary transmission routes, and that the primary source of information was from television (85.5%), and radio (56.1 %) [23].

**Table 6.** Level of Knowledge of the BHW towards preventing COVID-19 transmission(n=59)

| Knowledge | Correct |  | Wrong |  |
| --- | --- | --- | --- | --- |
|  | f | % | f | % |
| 1. The main clinical symptoms of COVID-19 are fever, loss of taste, fatigue, dry cough and body aches. | 59 | 100.00 | 0 | 0.00 |
| 2. Unlike the common cold, stuffy nose and sneezing are less common in persons affected with the COVID-19 virus. | 40 | 67.80 | 19 | 32.20 |
| 3. Currently, there is no effective cure for COVID-19, but early symptomatic and supportive treatment can help most patients recover from the infection. | 50 | 84.75 | 9 | 15.25 |
| 4. The vulnerable members of the population including the young, elderly, pregnant, and those with chronic illnesses are the ones likely to develop severe symptoms. | 51 | 86.44 | 8 | 13.56 |
| 5. RT-PCR is currently the accepted method to detect if a person has been infected by COVID-19 virus. | 57 | 96.61 | 2 | 3.39 |
| 6. Asymptomatic patients with COVID-19 infection are not infectious and cannot spread it to other people. | 44 | 74.58 | 15 | 25.42 |
| 7. A contact is a person who has face to face contact with a probable or confirmed case within 1 meter and for more than 15 minutes, or direct physical contact with a probable or confirmed case, or direct care for a patient with probable or confirmed COVID-19 disease without using proper PPE | 56 | 94.92 | 3 | 5.08 |
| 8. 3ply surgical masks are more efficient than cloth masks in preventing spread or acquiring the virus. | 56 | 94.92 | 3 | 5.08 |
| 9. Individuals with flu-like symptoms must be isolated in a separate room or area within the household | 58 | 98.31 | 1 | 1.69 |
| 10. It is not necessary for healthy individuals to take measures to prevent the spread of COVID-19 infection. | 48 | 81.36 | 11 | 18.64 |
| 11. To prevent the spread of COVID-19 infection, individuals should avoid crowded places and avoid taking public transportation. | 50 | 84.75 | 9 | 15.25 |
| 12. Any person below 21years old and above 60 years old, and those with immunodeficiency, comorbidity, or other health risks, and pregnant women including any person who resides with the aforementioned, shall be required to remain in their residence at all times, except when indispensable under the circumstance of obtaining essential goods and services, or for work industries and offices or other such permitted activities. | 57 | 96.61 | 2 | 3.39 |
| 13. I will encourage symptomatic individuals to stay at home unless there is a pressing need to go to a health facility for medical consultation. | 59 | 100.00 | 0 | 0.00 |
| Factor Average | 53 | 89.31 | 6 | 10.69 |

As observed in Figure 2, BHWs show that they are knowledgeable of protocols of isolation and quarantine. Although quarantine efforts are much in place, the correlation of higher number of cases of the selected barangays could be due to practical implications. As discussed in a study in Iloilo province, intrinsic motivation of staying alive can encourage a resident to follow quarantine protocol during the pandemic. However, these are also mediated by the need to maintain their livelihood [24]. Other factors are discussed from a study on How does age affect personal and social reactions to COVID-19: Results from the national Understanding America Study. It showed that once the spread of infection increases and after governmental advice on practicing preventive behaviors is phased in, older people may take more precautions. In addition, social desirability might have affected people’s response to preventive measures [25] .

However, there were relatively lesser degree of knowledge in the areas of “*Unlike the common cold, stuffy nose and sneezing are less common in persons affected with the COVID-19 virus*”, “*Asymptomatic patients with COVID-19 infection are not infectious and cannot spread it to other people*” and “*It is not necessary for healthy individuals to take measures to prevent the spread of COVID-19 infection*” which all got the least correct answers among all knowledge items. The lower awareness or gap in knowledge here can be correlated to the type of health care facility and professional care. As reported in a similar study in North Nigeria, those in primary and secondary health facilities had lower knowledge compared to those in tertiary hospitals, and that those in tertiary institutions were more involved in patient care and would seek more information compared to other staff [26]. This fraction of healthcare workers who are less knowledgeable could be a weak link and can increase the risk of contracting the disease [27].

Table 7 illustrates the general characteristics of the BHWs in terms of their knowledge where most of them were classified as knowledgeable (66.10%). None of them scored average to no knowledge at all which is a very good indication of the BHWs’ performance in terms of recognizing the different facts about COVID-19 prevention measures. This suggests that there is an adequate knowledge of the transmission of COVID-19, which is required to contain the virus through precautionary practices. This is also an indication that the DOH information drive plays a vital role in educating the BHWs about COVID-19. The factors that were assessed in this study include the age, sex, educational attainment, and monthly income of the participants. In previous studies, the aforementioned variables together with the occupation, religion, and the knowledge on the transmission and prevention of COVID-19 infection of the participant, are associated with the precautionary practices towards COVID-19 transmission. The results of this study showed no significant difference between the knowledge score of the participants. This could be “attributed to the fact that the proponents of this study are targeted only to the BHWs, majority of which are women and in the same income bracket.

**Table 7.** The Distribution of the Respondents based on their Level of Knowledge (n=59)

| Score | Knowledge Level | f | % |
| --- | --- | --- | --- |
| 13 | Very Knowledgeable | 20 | 33.90 |
| 9 to 12 | Knowledgeable | 39 | 66.10 |
| 5 to 8 | Average | 0 | 0.00 |
| 1 to 4 | Less Knowledgeable | 0 | 0.00 |
| 0 | No Knowledge At All | 0 | 0.00 |

The participants were found to have good levels of knowledge regarding COVID-19 transmission. This is expected considering the line of work that they are in. BHWs are the first line of contact between the health care system and the community. As such, they are expected to provide basic health education and primary health care services and link patients to appropriate health care facilities, as necessary. A Philippine study on KAP of COVID-19 among income-poor households showed that a significant proportion (49.4%) of the study population said they would contact a BHW should they experience symptoms such as fever, cough or sore throat, signifying the importance of BHWs in the participants’ health-seeking intentions [28].

Future behavior is shown to be affected by assessing knowledge of precautionary measures in contracting the disease[29]. A study on KAP towards COVID-19 among Chinese residents showed that higher COVID-19 knowledge scores were found to be significantly associated with a lower likelihood of negative attitudes and potentially dangerous practices towards COVID-19, indicating the importance of health education among the participants [30]. Previous studies also reported that the knowledge about an infectious disease significantly affects the behavior of the individuals in order to prevent infection.

Table 8 shows the general attitude of the BHWs, a factor average of 4.15 was computed which was translated as favorable. This rating was attributable to the high perceptions on the items “*I make it a habit to frequently wash my hands for 20 seconds with soap or hand sanitizer*” and “*I promote proper nutrition, and hydration to increase the body’s immunity and resistance to COVID-19*” both with 4.68. Likewise, the respondents also identified very favorable treatment to the idea that “*I constantly remind people the importance of wearing masks, face shields and social distancing*” with 4.66.

**Table 8.** Type of Attitude of the BHWs towards preventing COVID-19 transmission (n=59)

| Attitude | Weighted Mean | Standard Deviation | Description | Kind of Attitude |
| --- | --- | --- | --- | --- |
| 1. The imposition of wearing a face mask, face shield, and social distancing is a violation of the constitutional right to liberty and about making decisions about own health and bodily integrity. | 3.05 | 1.64 | Neutral | Neutral |
| 2. I constantly remind people the importance of wearing masks, face shields and social distancing | 4.66 | 0.66 | Strongly Agree | Very Favorable |
| 3. I make it a habit to frequently wash my hands for 20 seconds with soap or hand sanitizer. | 4.68 | 0.47 | Strongly Agree | Very Favorable |
| 4. I would avoid public places as much as possible except when I need to buy essentials | 4.59 | 0.50 | Strongly Agree | Very Favorable |
| 5. I would compromise family/friend gatherings in order to observe the social distancing protocols | 4.44 | 0.68 | Strongly Agree | Very Favorable |
| 6. Social distancing and wearing of face masks ends when I arrive home, even though I have working age group members in the household | 3.15 | 1.28 | Neutral | Neutral |
| 7. I constantly update myself with news such as about the new COVID-19 strain. | 4.58 | 0.59 | Strongly Agree | Very Favorable |
| 8. I correct people about misinformation with COVID-19 such as gargling salty water can prevent infection | 3.49 | 1.29 | Agree | Favorable |
| 9. I prefer virtual meetings if it is not extremely necessary to meet face to face. | 4.20 | 0.78 | Strongly Agree | Very Favorable |
| 10. I promote proper nutrition, and hydration to increase the body's immunity and resistance to COVID-19 | 4.68 | 0.47 | Strongly Agree | Very Favorable |
| 11. I will not hesitate to get the COVID-19 vaccine. | 4.12 | 1.00 | Agree | Favorable |
| **Factor Average** | **4.15** | **0.85** | **Agree** | **Favorable** |

Legend: 1.00-1.79 -Very Unfavorable, 1.80-2.59 -Unfavorable, 2.60-3.39-Moderate, 3.40-4.19-Favorable, 4.20-5.00-Very Favorable

The BHWs on the other hand were in neutral state when asked about “The imposition of wearing a face mask, face shield, and social distancing is a violation of the constitutional right to liberty and about making decisions about own health and bodily integrity” and “Social distancing and wearing of face masks ends when I arrive home, even though I have working age group members in the household”.

Overall, the results show a general willingness for BHWs to make behavioural changes in the face of the COVID-19 pandemic. While it showed that knowledge didn’t necessarily affect the BHWs attitude towards COVID-19 (Table 5), these results may be attributed to higher knowledge scores among them, in which 66.10% were classified as knowledgeable. This is reinforced in other studies in which high levels of attitudes were also seen in a study conducted in China and in South Korea, showing that respondents with higher knowledge displayed positive attitudes such as wearing masks and practicing hand hygiene [30]. Another reason may be attributable to the response strategy made by the national government via DOH which includes community engagement activities to promote preventive behaviors [31]. This is also reflected in a study conducted by Azlan et al. which showed that those working in the public sector have the highest positive attitude toward COVID-19[57].

The neutral response on statement no. 1 may suggest that respondents are less inclined to express their opinion regarding the subject matter of constitutional rights. The COVID-19 pandemic has become a public threat that could justify restrictions on certain rights such as mentioned in statement no. 1. According to the United Nations, all States have a duty to protect human life, including by addressing the general conditions in society that give rise to direct threats to life [32]. Adopting extraordinary measures such as wearing of face masks, face shields, and social distancing preserves life that citizens must abide for the common good. The neutral response on statement no. 6 on the other hand, may be attributed to the respondents’ lesser degree of knowledge in the areas regarding asymptomatic and healthy individuals (Table 2). This may reflect a need to address these gaps in knowledge for BHWs to better understand COVID-19 and how it is transmitted, as this is key to form more positive attitudes towards effectively fighting the disease [33].

In Table 9, the BHWs generally recorded a practice classified as Always with an average of 4.43 in terms of practice. This can be associated with the fact that they “*clean their hands with alcohol-based sanitizers when soap and water is not available*” (4.76), *“wash their cloth mask at least daily*” (4.75), “*wear face masks/face shields properly when they go out for work/errands* “(4.75) and “*discard their disposable face masks and other PPE in the proper waste bin after use*”.

**Table 9.** Extent of Practices of the BHWs towards preventing COVID-19 transmission(n=59)

| Practices | Weighted Mean | Standard Deviation | Extent of Practice |
| --- | --- | --- | --- |
| 1. Do you practice frequent and proper handwashing with soap and water for at least 20 seconds before and after getting into contact with other people or high-touch objects/surfaces? | 4.51 | 0.77 | Always |
| 2. Do you clean your hands with alcohol-based sanitizers when soap and water is not available? | 4.76 | 0.47 | Always |
| 3. Do you practice proper cough etiquette? | 4.71 | 0.72 | Always |
| 4. Do you practice social distancing protocols when you interact with your coworker/s or other individual/s? | 4.42 | 0.72 | Always |
| 5. Do you clean high-touch objects/surfaces with disinfectants regularly? | 4.34 | 0.78 | Always |
| 6. Do you wear face masks/face shields properly when you go out for work/errands? | 4.75 | 0.48 | Always |
| 7. Do you wear face masks/face shields properly when you interact with your coworker/s or other individuals/s? | 4.66 | 0.58 | Always |
| 8. Do you wash or sanitize your hands before you put on your face masks/face shields? | 4.34 | 0.76 | Always |
| 9. Do you discard your disposable face masks and other PPE in the proper waste bin after use? | 4.75 | 0.48 | Always |
| 10. Do you avoid going to public gatherings or other crowded areas such as malls and supermarkets? | 4.32 | 0.84 | Always |
| 11. Do you avoid getting into contact with persons experiencing respiratory symptoms? | 4.09 | 1.06 | Often |
| 12. Do you avoid getting into contact with persons who are at high-risk for severe COVID-19 such as those who have comorbidities and are immunocompromised? | 4.19 | 1.11 | Often |
| 13. Do you stay at home to isolate when it is your off-day from work? | 4.32 | 0.82 | Always |
| 14. Do you avoid eating outside with other people such as your coworker/s? | 3.93 | 0.87 | Often |
| 15. Do you avoid using public transportation as a means to go to work? | 3.73 | 1.05 | Often |
| 16. Do you monitor your health? | 4.71 | 0.62 | Always |
| 17. Do you monitor the health of the people around you? | 4.22 | 1.00 | Always |
| 18. Do you persuade your coworker/s and other individual/s to follow the safety protocols implemented by your local government? | 4.59 | 0.65 | Always |
| 19. Do you wash your cloth mask at least daily? | 4.75 | 0.66 | Always |
| **Factor Average** | **4.43** | **0.76** | **Always** |

Legend: 1.00-1.79 -Never, 1.80-2.59 -Rarely, 2.60-3.39-Sometimes, 3.40-4.19-Often, 4.20-5.00-Always

As to the areas that they were not accustomed to practicing were more on “avoiding the use of public transportation as a means to go to work” and “avoiding eating outside with other people such as your coworker/s”.

In this study, the BHW’s practices were found to be mostly appropriate. However, factors like transportation and certain living circumstances contribute to some of the poor practices seen. In a study in China, it was shown that 89.7% of the surveyed HCWs followed correct practices regarding COVID-19, showing that practices are associated with work experience, working time, and other factors [34].

Other than the original duties and responsibilities of BHWs in the Philippines, during the COVID-19 pandemic, they are responsible to: identify persons who have symptoms of COVID-19; track families who are in quarantine; and monitor and care for patients in community isolation units. Their work experience exposed them to patients who are probable, suspected or confirmed with COVID-19, thus, they may be infected with the virus if they are not careful. To prevent the transmission of the virus, Guidelines on Local Isolation and General Treatment Areas for COVID-19 cases (LIGTAS COVID) provided steps on proper hand hygiene, respiratory etiquette, mask management, and protocol for PPE use for each staff of LIGTAS COVID Center to be followed [35]. These may explain why the respondents had *Always* as the extent of practice for “*wash their cloth mask at least daily*”, and “*discard their disposable face masks and other PPE in the proper waste bin after use*”.

Part of the duties of BHWs is to educate the community about COVID-19 prevention. Their practices on “*wearing face masks/face shields properly when they go out for work/errands*” may be explained by their very favorable attitude to the idea that “*I constantly remind people the importance of wearing masks, face shields and social distancing*”. Their practice of “*clean their hands with alcohol-based sanitizers when soap and water is not available*” may be associated with their high perceptions on “*I make it a habit to frequently wash my hands for 20 seconds with soap or hand sanitizer*”.

They are not accustomed to “avoiding the use of public transportation as a means to go to work” and this may be associated with their lesser knowledge on “to prevent the spread of COVID-19 infection, individuals should avoid crowded places and avoid taking public transportation”.

Using Pearson rho-correlation analysis, Table 10 shows that knowledge does not necessarily affect attitude (p-value >0.05) but significantly contributes to improving attitude and frequency of practice among the BHWs. Likewise, attitude significantly (p-value < 0.05) induced a positive impact (coefficient =0.34) on the practices, that is, as the attitudes of the respondents were more favorable, the more frequent the practices were in preventing COVID-19 transmission.

**Table 10.**
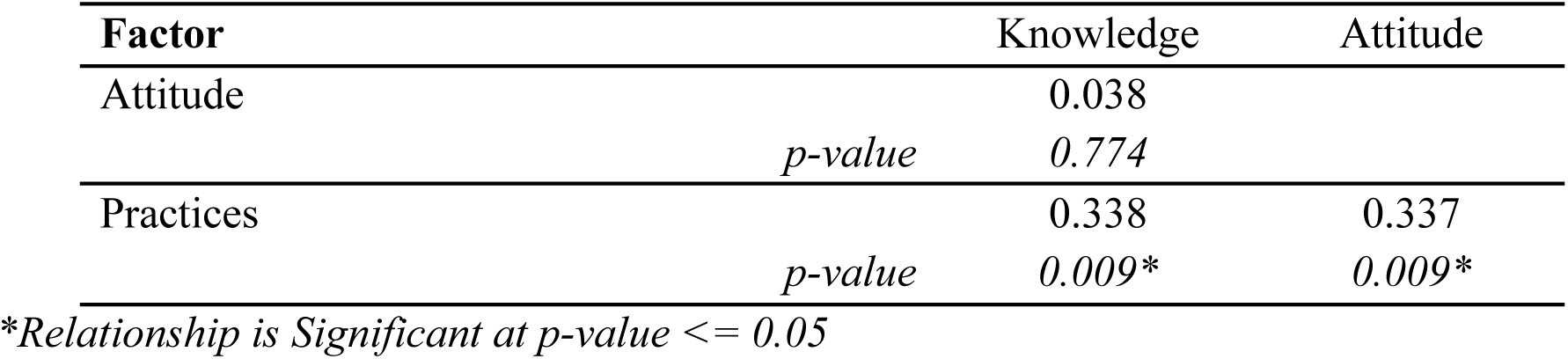
The Relationship Between the Factors used in the Study.

| **Factor** | Knowledge | Attitude |
| --- | --- | --- |
| Attitude | 0.038 |  |
| *p-value* | 0.774 |  |
| Practices | 0.338 | 0.337 |
| *p-value* | 0.009* | 0.009* |

\*Relationship is Significant at $p\text{-value} \leq 0.05$

Table 11 shows the effect size of the knowledge and type of attitude of the respondents on the different measures to combat the transmission of the COVID-19. It can be shown that a unit increase in knowledge increases the frequency of practice by 0.7 units and every unit increase of attitude adds up to contributing 0.21 to the frequency of practices.

**Table 11.**
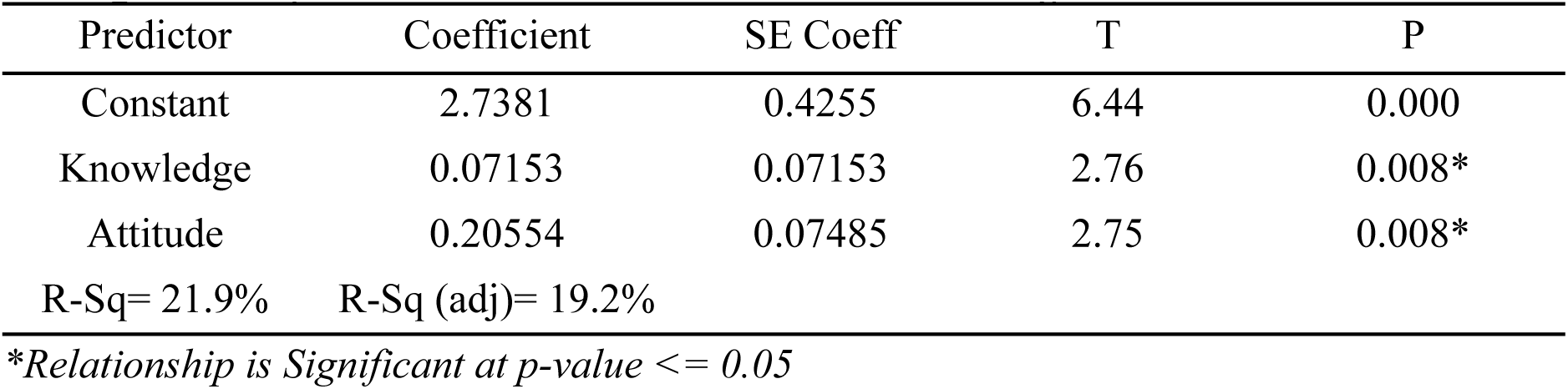
Regression Analysis: Practices versus Knowledge, Attitude. The regression equation is Practices = 2.74 + 0.0715 ***Knowledge*** + 0.206 ***Attitude***

The r-square value (coefficient of variation) also indicates that 19.2%-21.9% of the total variations of practice can be explained by the variations of knowledge and attitude. The other explanatory factors are external to the current study.

## CONCLUSION

This study examined KAP of BHWs from the top 5 barangays in Cebu City with the highest COVID-19 cases. Among the 59 respondents, all were females, and majority were highschool graduates, around 49-58 years of age average. This study found that the participants were knowledgeable about COVID-19 transmission, particularly of the main clinical symptoms and the protocols of isolation and quarantine. The BHWs’ knowledge showed significant improvement in attitude and frequency of practices. This association between knowledge and practices demonstrates the importance of prompt and accurate information regarding COVID-19. The general attitude of the BHWs were favorable which significantly induces a positive impact on practices on preventing transmission – use and disposal of PPEs, hand washing/sanitizing. However, there were some reservations regarding social distancing and wearing of face masks at home. Furthermore, they were not accustomed to avoiding public transportation and eating outside with other people which probably correlates with certain factors and living circumstances. Therefore, COVID-19 prevention strategies should be employed to address these specific gaps in practices.

## RECOMMENDATIONS

The proponents would like to recommend the validation of the questionnaire by public health/family and community medicine experts with regards to its formulation and relevance.

It is also recommended that health education programs should be reinforced to narrow the gap in knowledge among BHWs. With this, an increase in knowledge, attitudes, and practices among them improve. This could also affect their perception towards the significance of the study, as it is noted that there was difficulty in gaining their participation which may be due to their lack of understanding.

The majority of income-poor households go to BHWs whenever they experience symptoms such as fever, cough, or sore throat, therefore, primary health care must also be given attention similar to secondary and tertiary hospitals. If patients are being taken care of at the primary level, this prevents them from going to secondary and tertiary hospitals unless for referrals. This would then provide more room for tertiary hospitals to accommodate severe cases of COVID-19 infection.

Another point of interest would be to assess the KAP of the general public since it could give an even better understanding with regards to the status of KAP and factors that affect the former. This could provide an insight as to which socio-demographic profile could significantly affect the KAP that could help predict the future response of the public to infectious disease outbreaks.

## Data Availability

All data produced in the present work are contained in the manuscript

## ACKNOWLEDGEMENT

The accomplishment of this research would not have been made possible without the assistance, support, and guidance of the following:

To God Almighty, who provided the researchers with the strength, wisdom, good health and blessing needed to finish this research paper.

To our parents, who supported us throughout this year despite the difficulties brought about by the pandemic.

To all the BHWs, for their voluntary participation in the study. We highly appreciate their time and effort to answer the survey despite the limitations brought about by the COVID-19 pandemic. To Dr. Jeffrey B. Ibones, the head of the City Health office for granting us permission to proceed with our study.

To Mr. Mark Borres, the statistician, for helping us analyze our data. To Mr. Jules Barry Pabenal for proofreading our paper. To Mr. Abdul Rauf Sissay for helping us translate our questionnaires to the Visayan dialect.

To Cebu Institute of Medicine, for allowing us to conduct this research considering that our target population is outside the school premises.

To group 7, for helping each other in this research and for supporting each other through this academic year.

Lastly, to Dr. Maria Philina P. Villamor, our research adviser, for her guidance throughout the whole research process, and for allotting her time to check and review our paper and providing feedback and comments that will help us improve our paper.

## Appendix A. PARTICIPANT INFORMED CONSENT FORM

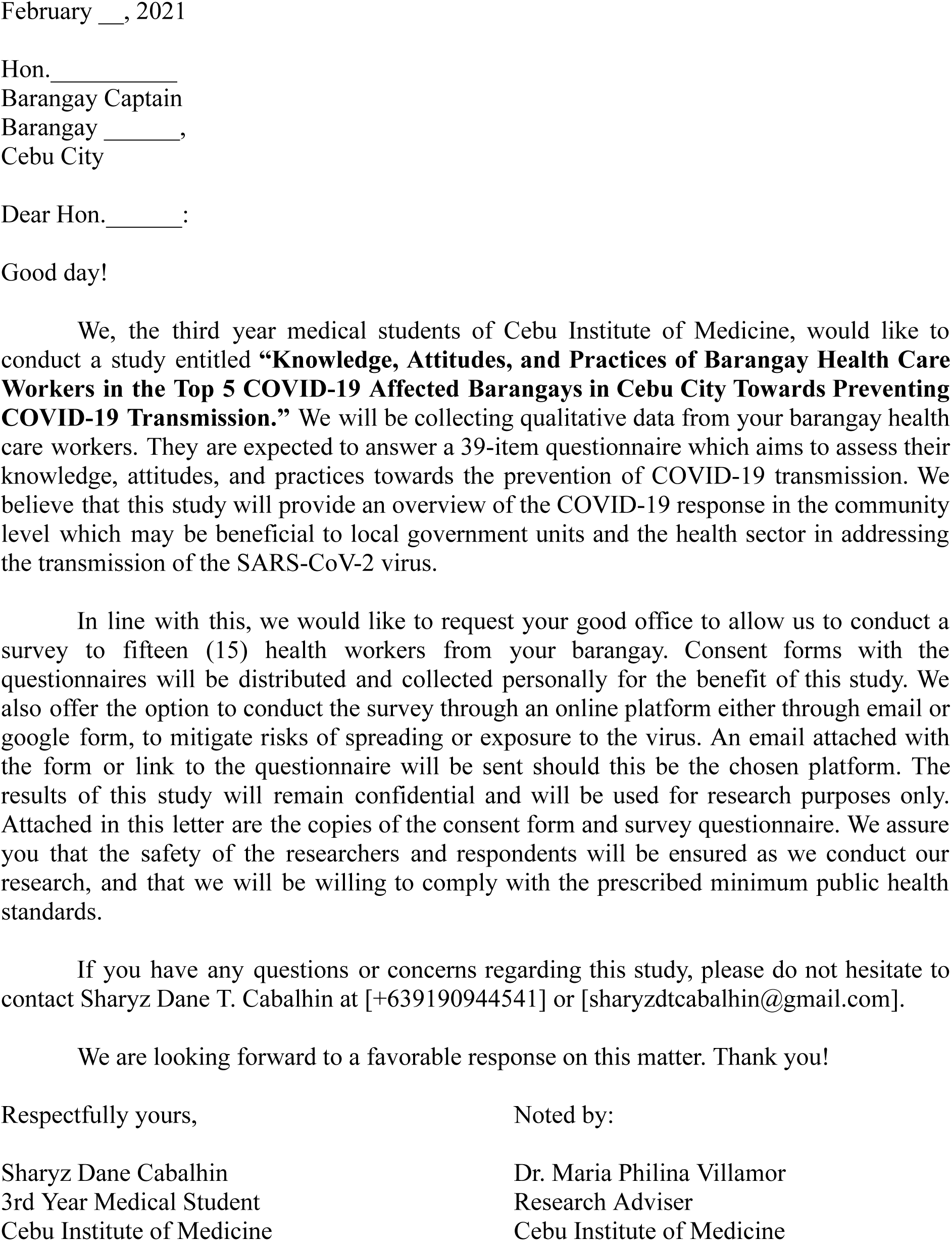

## Appendix B. RESEARCH QUESTIONNAIRE

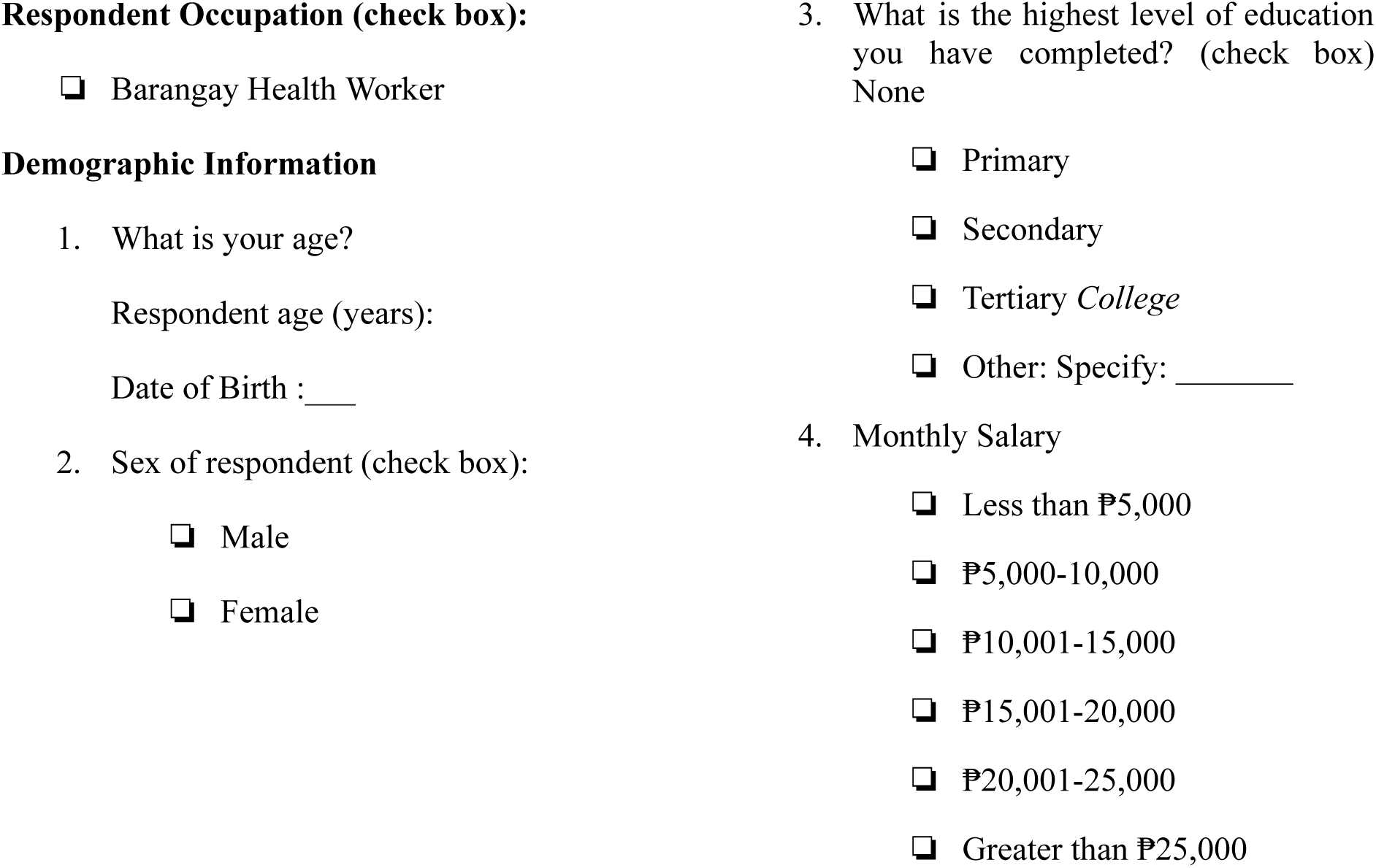

**Knowledge**

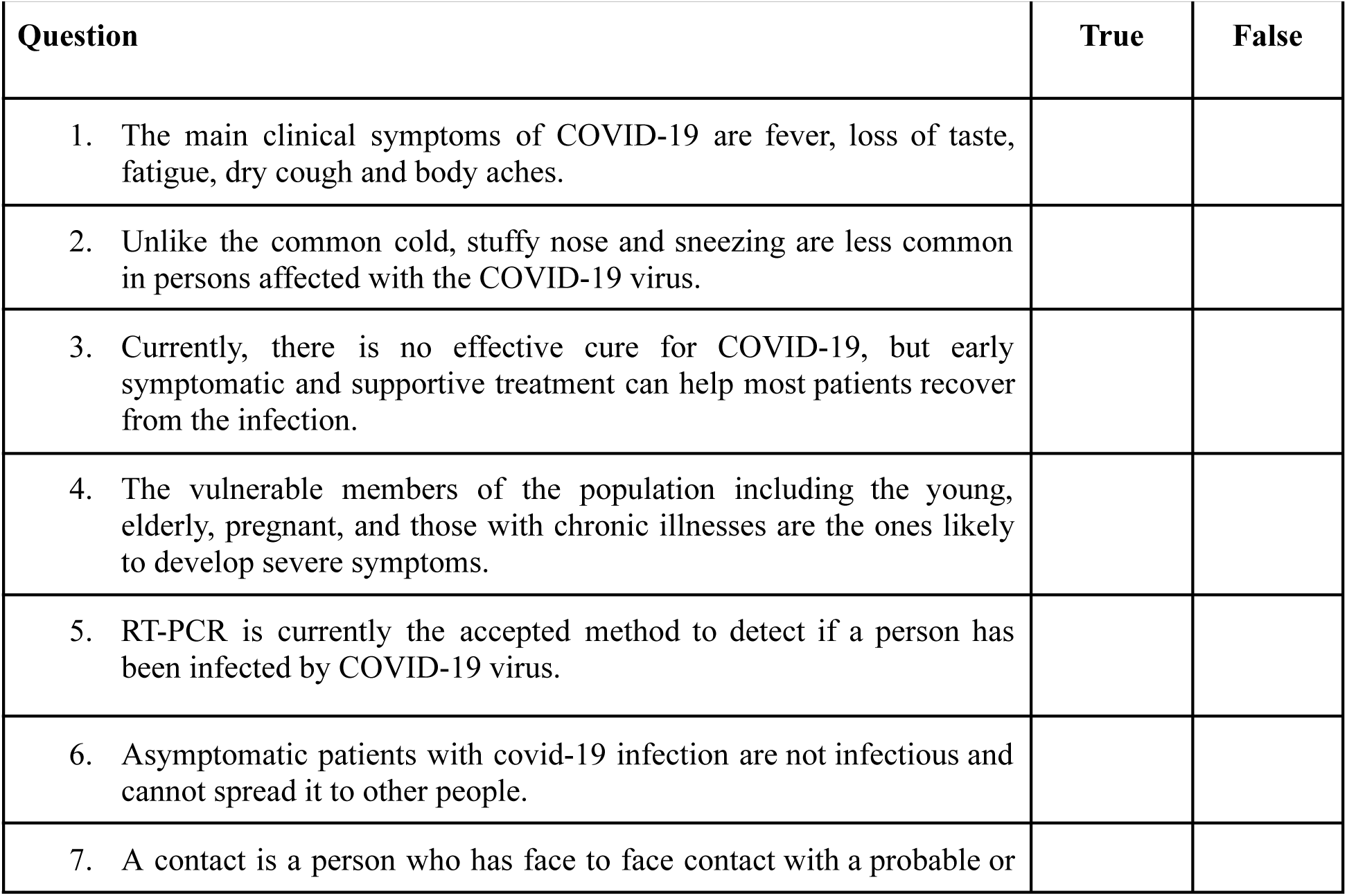
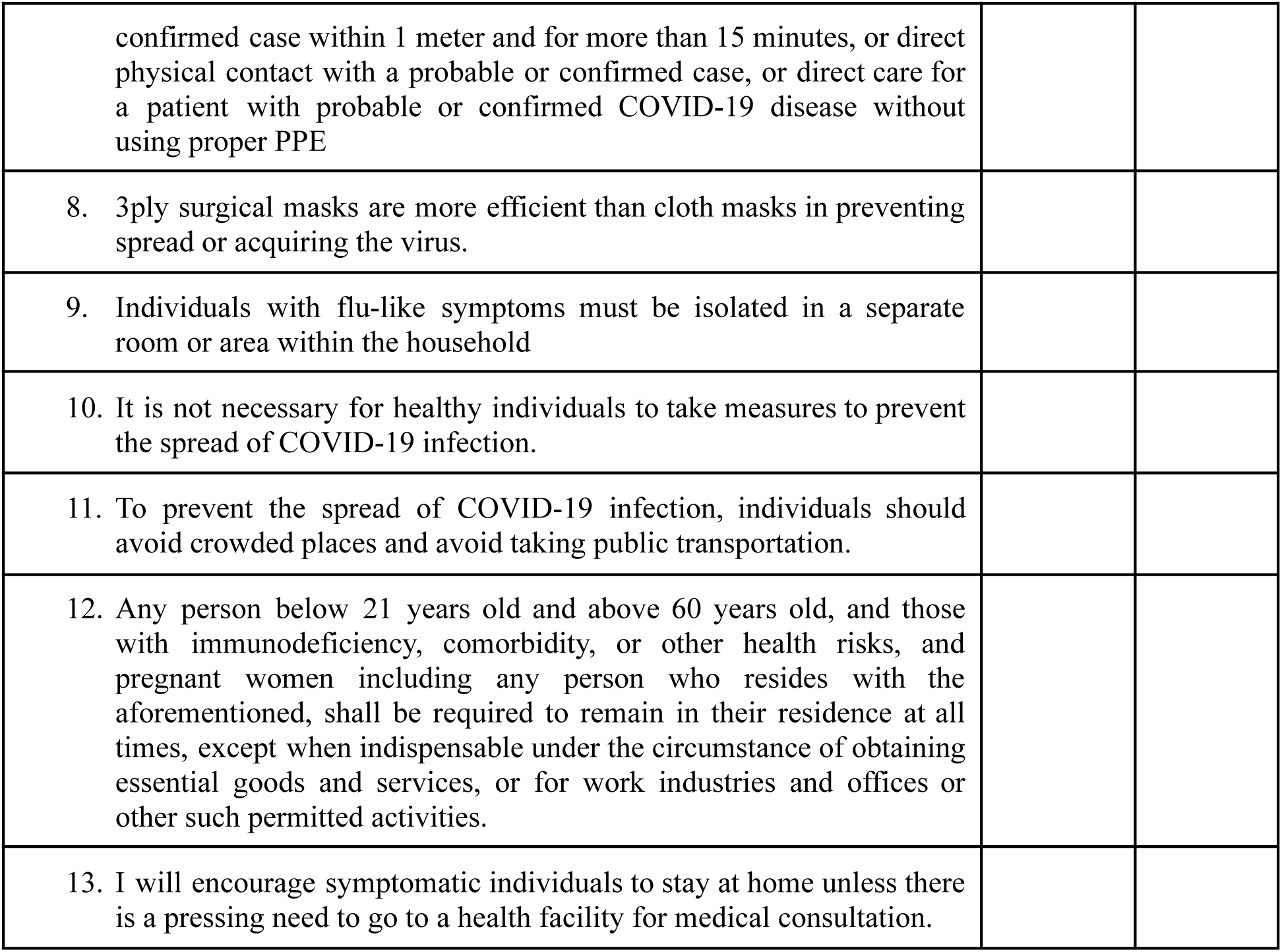

**Attitude**

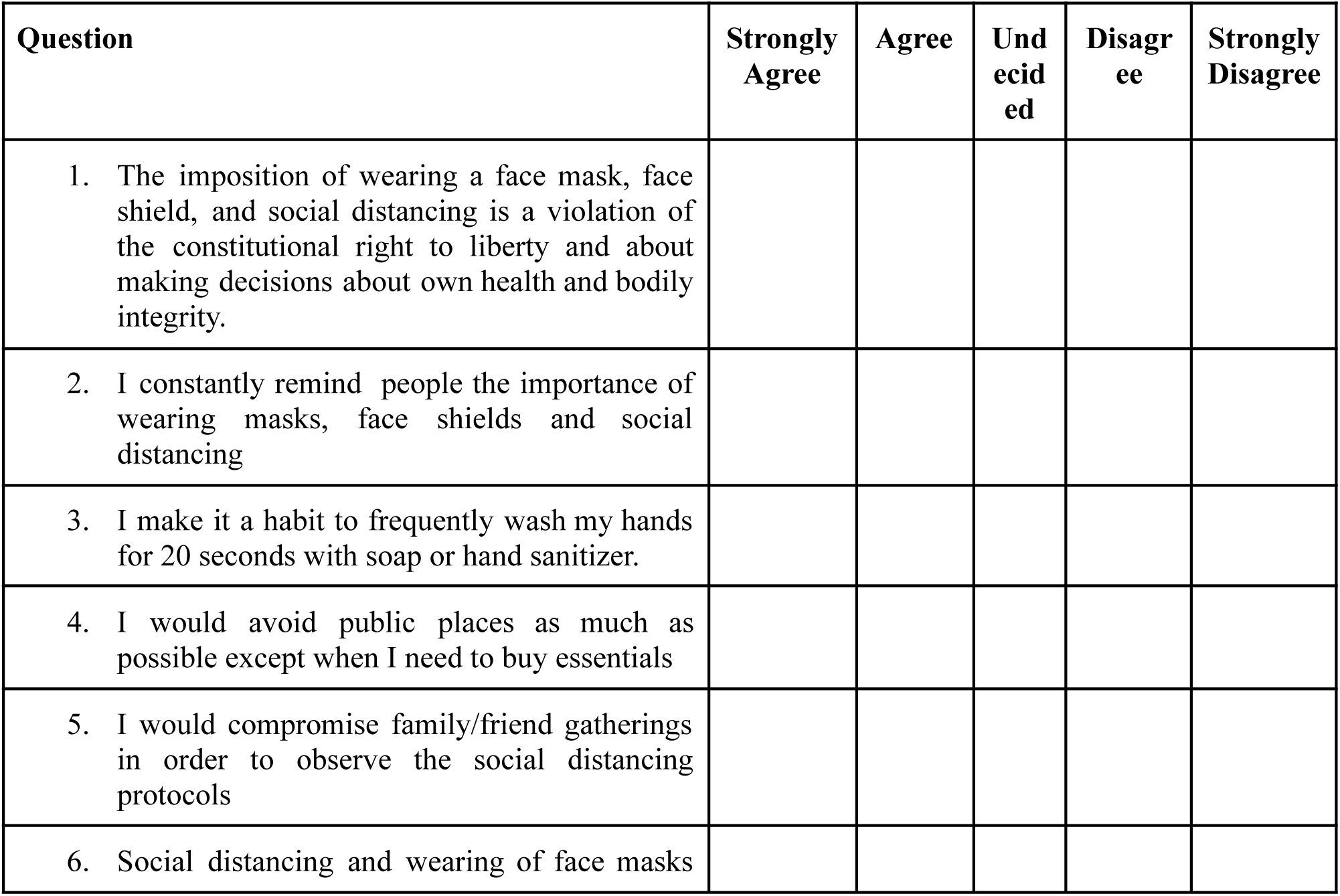
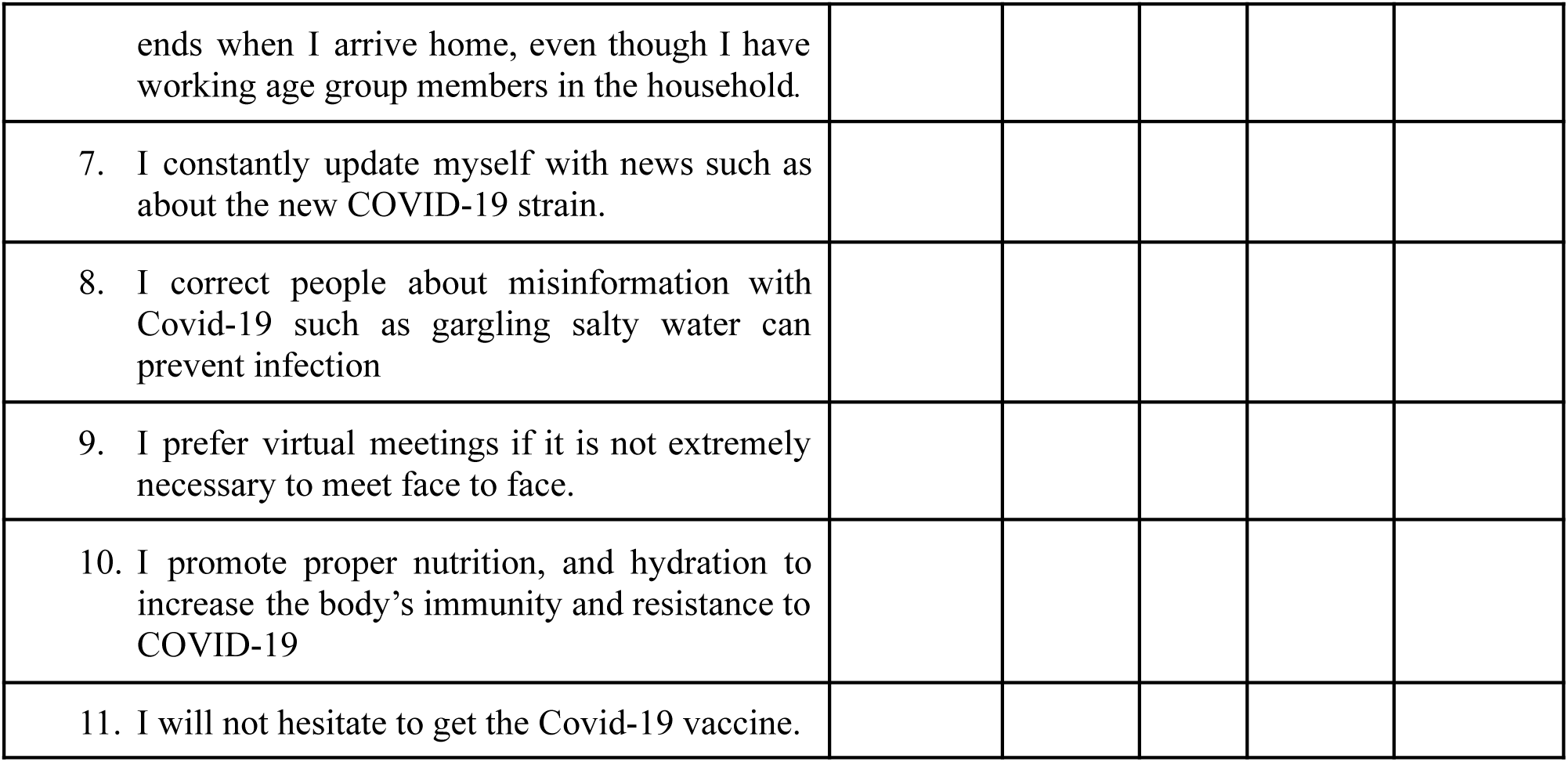

**Practices**

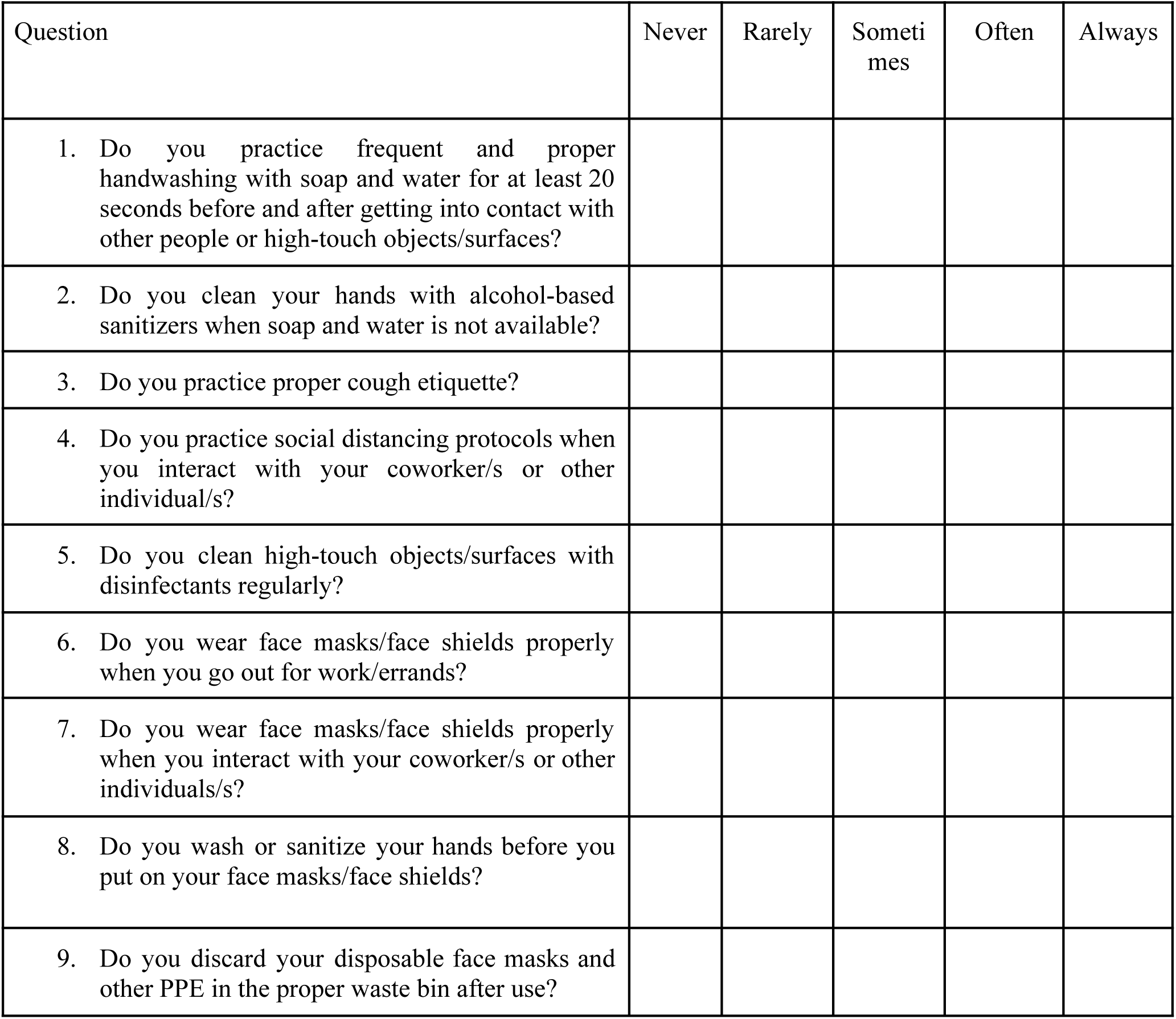
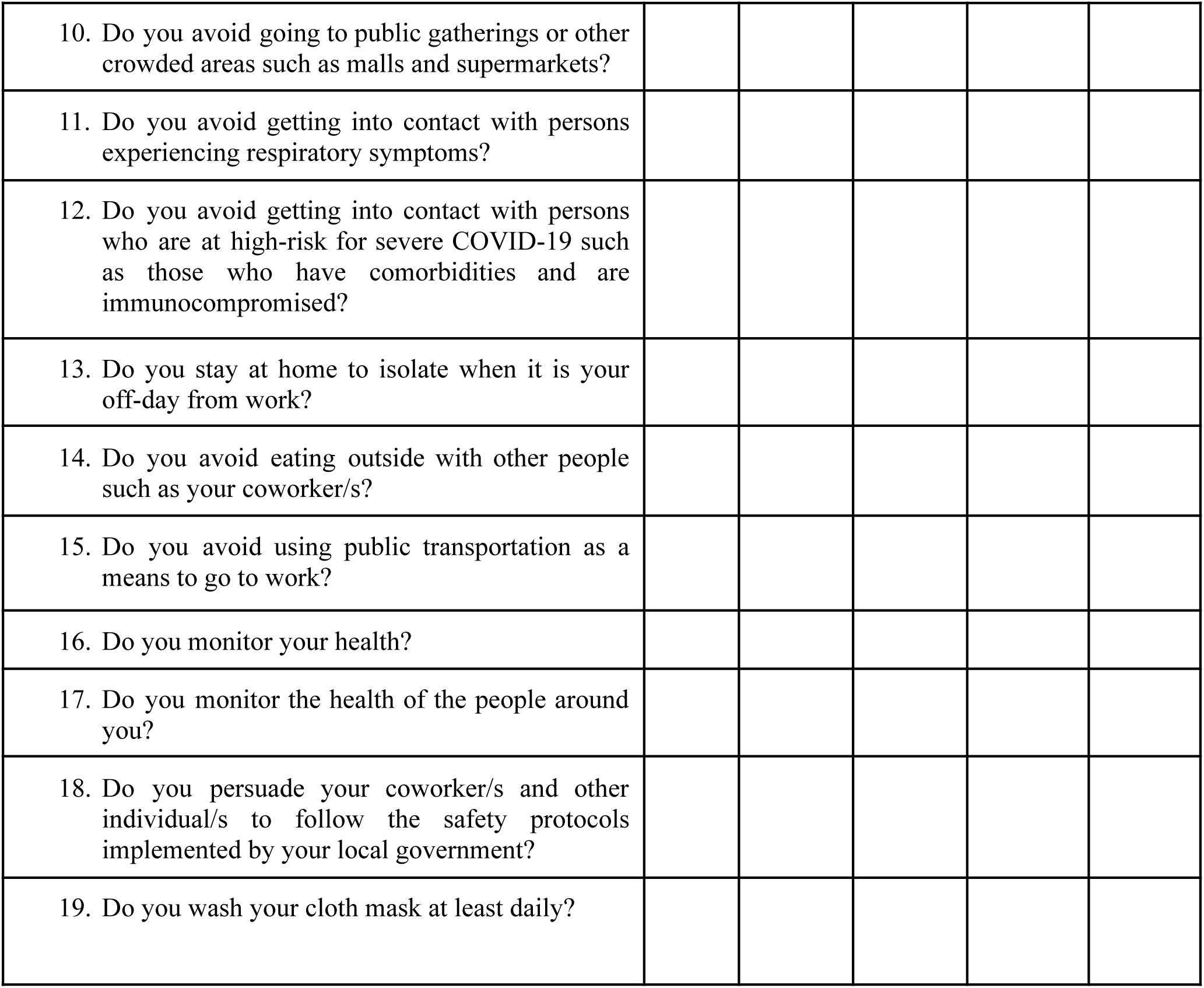

## Appendix C. Letter to the City Health Officer

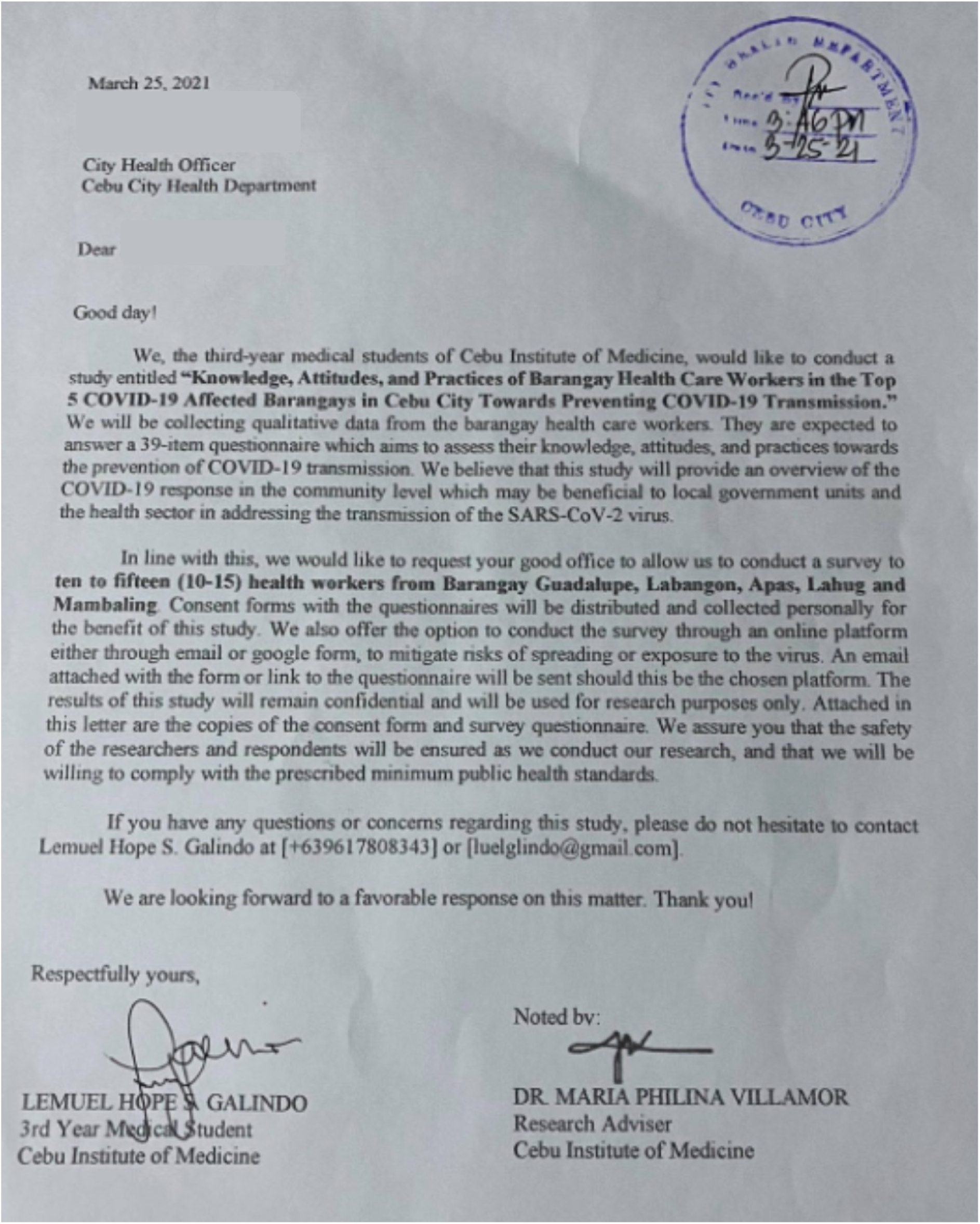

## Appendix D. Sample Letter to the Barangay Captains

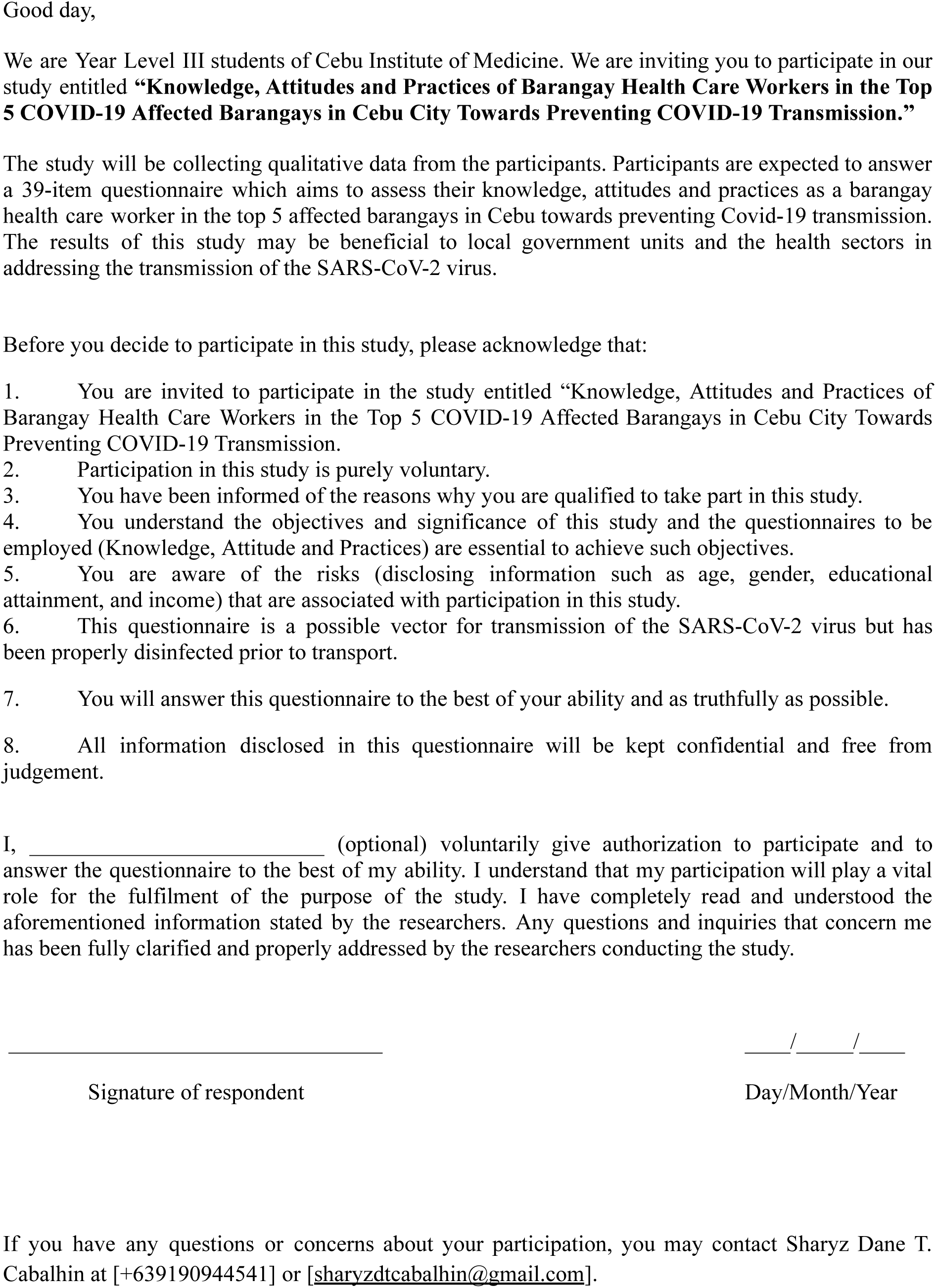

## Appendix E. BUDGET

| **Specifics** | **Unit cost** | **Quantity** | **Total Cost** |
| --- | --- | --- | --- |
| Questionnaire printing | 7.00 | 84 | 588.00 |
| Fuel Cost | 500.00 | 3 | 1,500.00 |
| **TOTAL:** |  |  | **2,088.00** |

## Appendix F. Gantt Chart

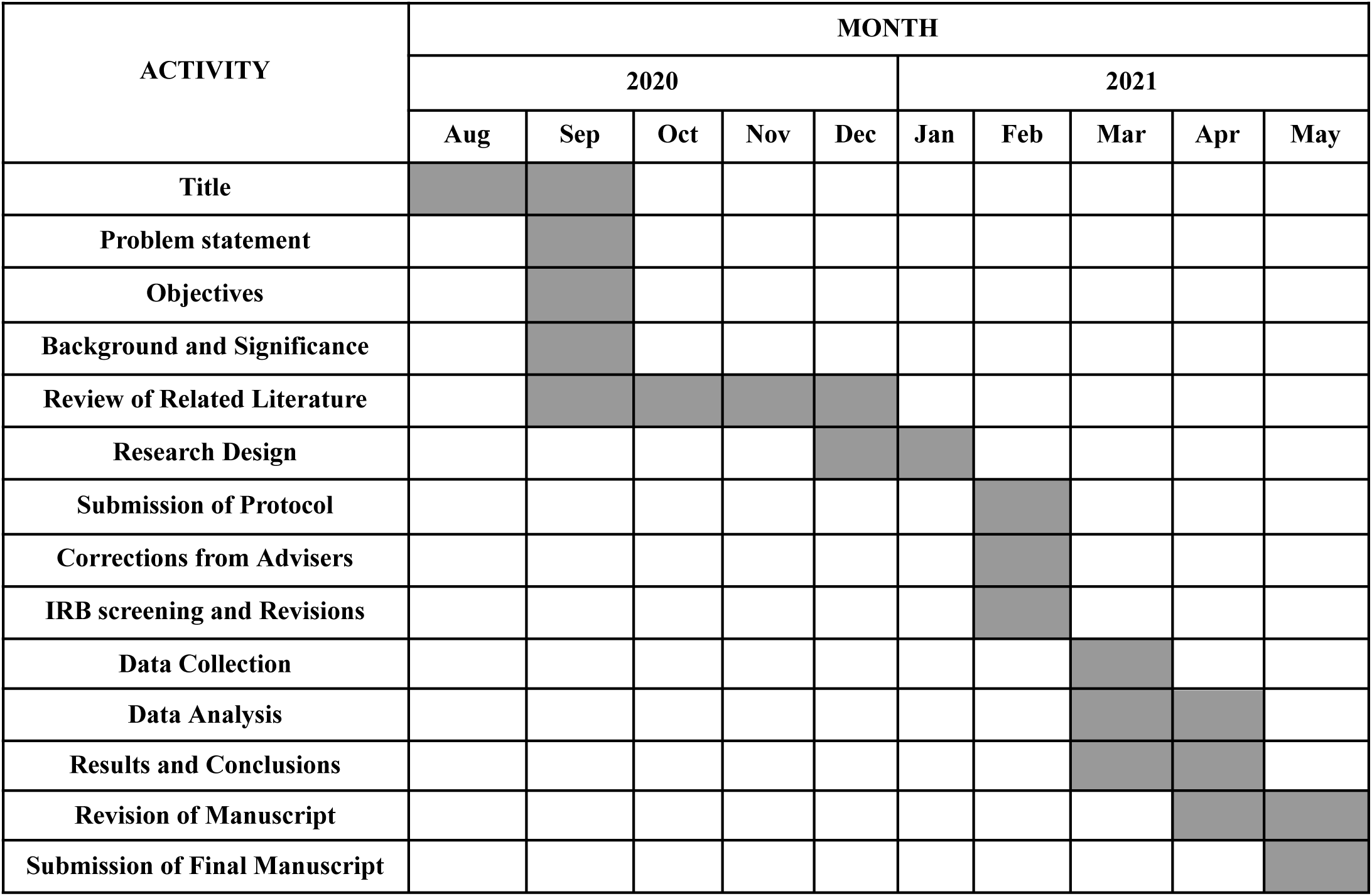

